# Six-Week Changes in Pain Biomarkers Following Reverse Total Shoulder Arthroplasty: A Prospective Cohort Study

**DOI:** 10.64898/2026.02.10.26346010

**Authors:** Chris J. Pierson, Andrew Nasr, Cassie M. Argenbright, Bansari Thakkar, Alison Cabrera, Tracy L. Greer, Khosrow Behbehani, Robin Jarrett, Jason Zafereo

**Author notes:** **Corresponding Author:** Chris J. Pierson Current, Chris J. Pierson is currently a postdoctoral fellow at the University of Texas Medical Branch.

## Abstract

**Background:** Reverse total shoulder arthroplasty (rTSA) is an increasingly common surgical procedure often performed to treat pain related to glenohumeral osteoarthritis or to rotator cuff arthropathy. Although surgical outcomes are generally excellent, recent evidence has found that postoperative pain (≥ 3/10) two years following surgery is reported by an estimated 18% of patients. Recently, the NIH Acute-to-Chronic Pain Signatures program recommended longitudinal studies using select biomarkers to describe and predict individual patient responses to surgery. These data are not yet available for rTSA procedures.

**Methods:** This was a longitudinal cohort study performed at a single academic medical center. Twenty participants undergoing rTSA surgery were included, recruited from a tertiary hospital system in the southern United States. The first objective of this study was to describe changes in general pain intensity (Numerical Pain Rating Scale), widespread body pain, anxiety (General Anxiety Disorder-7), depression (Patient Health Questionnaire-9), neuropathic pain symptoms (painDETECT), and quantitative sensory testing from baseline to 6 weeks following rTSA. The second objective was to identify the baseline demographic and pain-related factors associated with 6-week postsurgical improvements in pain intensity.

**Results:** From before to after surgery, our cohort demonstrated significant improvement in shoulder pain intensity, widespread body pain, PainDETECT score, and temporal summation magnitude measured at the surgical deltoid. Degree of 6-week pain intensity improvement was associated with baseline pain intensity (F=18.79, p=0.0004) and temporal summation magnitude of the tibialis anterior (F=5.06, p=0.0380).

**Conclusions:** Pain intensity, location, nature, and mechanism can serve as biomarkers of the short-term postsurgical changes that can be expected following rTSA. Baseline pain intensity and temporal summation magnitude of the tibialis anterior were associated with the degree of pain improvement, suggesting their use for preoperative risk assessment. Future research should evaluate whether these 6-week biomarker changes are associated with the development of chronic postoperative pain at longer durations after surgery.

**Level of Evidence:** Level I, Prognostic Study

## Introduction

Reverse total shoulder arthroplasty (rTSA) is an increasingly common surgical procedure used to treat a wide range of degenerative glenohumeral diseases and irreparable rotator cuff arthropathies.^1^ From 2011 to 2017, the number of rTSA procedures performed in the United States alone increased from 22,835 to 62,705.^2^ Although surgical outcomes are generally excellent,^3^ recent cohort-level evidence has found that persistent postoperative pain (≥ 3/10) two years following surgery is reported by an estimated 18% of patients and is predicted by several preoperative factors, including baseline pain intensity.^4^ A small proportion of patients reporting persistent pain can be explained by surgical complications, hardware complications, or other structural pathologies. An overview of 1,259 rTSA cases from 2006 to 2016 found that ten reported an infection (0.8%), fourteen had a postoperative acromion or spine fracture (1.1%), eleven had a periprosthetic fracture (0.9%), and fifteen demonstrated prosthetic instability or dislocation (1.2%). Other rare complications exist, but each accounted for under 0.5%.^5^ With the 4% prevalence of structural complications being far lower than the 18% prevalence of persistent pain, non-structural pain mechanisms likely contribute to persistent pain for a significant number of patients.

The current need for research investigating the development of persistent post-operative pain is underscored by recent funding from the National Institutes of Health Common Fund for the Acute-to-Chronic Pain Signatures Consortium (A2CPS). This initiative recommends studying a select set of biomarkers for both the prediction of outcomes and the measurement of response to surgery. The five outcome measure categories recommended are patient-reported outcome measures (PROs), patient characteristics, -omics, quantitative sensory testing, and brain imaging.^6^

Surgical outcome prediction research has been performed in patients undergoing total knee arthroplasty (TKA)^7,8^ but not yet for rTSA. In short-term studies of patients undergoing TKA, reduced pressure pain threshold (PPT) prior to surgery on the arm (control site) was predictive of moderate/severe pain 24 hours following TKA.^7^ In another study, one-year PRO scores were predicted by arm but not knee PPT.^8^ In a combined cohort of 254 patients undergoing total hip arthroplasty (THA) and 239 undergoing TKA, preoperative pain sensitivity measured with PPT at the forearm was significantly associated with higher preoperative pain severity. Lower preoperative PPTs were associated with higher pain severity 12 months following THA, but the magnitude of pain improvement was not predicted by baseline PPT in either TKA or THA.^9^ The breadth of these studies highlights the significant gap in our understanding of rTSA outcome prediction, especially compared to lower extremity arthroplasty surgery.

To address this gap, the first objective of this study was to perform a descriptive analysis of 6-week outcome measure changes in a cohort of twenty patients who underwent rTSA. Based on results of TKA studies, we hypothesized that resting pain intensity, anxiety, depression, neuropathic pain symptoms, and surgical site pain sensitivity would demonstrate clinically significant improvements 6-weeks following rTSA. The second objective was to identify baseline demographic and pain-related predictive factors that were associated with greater 6-week pain improvements. We hypothesized that younger age, lower measures of central sensitization, and lesser psychological impairment would be associated with greater 6-week composite pain improvement.

## Methods

### Study Design

This was a single-group longitudinal cohort study from the Reverse Total Shoulder Arthroplasty: Postoperative Recovery & Outcomes (RTSA-PRO) cohort. All participants underwent a similar rTSA procedure at a single orthopaedic surgery center by two fellowship-trained shoulder surgeons.

### Setting

This study was approved by the Institutional Review Board at the University of Texas (UT) Southwestern Medical Center. Study data were collected and managed using REDCap electronic data capture tools hosted at UT Southwestern Medical Center.^10,11^ All participants were recruited from the UT Southwestern Orthopaedic Surgery Shoulder Clinic in Dallas, Texas. Enrollment began on June 23, 2023, and final data collection was completed on July 23, 2025.

### Participants

Included participants were over the age of 18, had radiographic evidence and a physician diagnosis of glenohumeral osteoarthritis or rotator cuff arthropathy, had pain for a duration of at least three months, and had completed two data collection visits. Exclusion criteria were signs of fracture or infection, substantial non-shoulder causes of pain, inability to read or write in English, a current central or peripheral neurologic condition affecting the ipsilateral shoulder, impaired sensation to light touch, trauma to the prosthesis following surgery, and pregnancy.

Participant recruitment was performed by a clinical treatment team of surgeons and physical therapists. All eligible participants were approached consecutively for recruitment. All data were collected by a licensed physical therapist in a quiet clinical laboratory or clinical examination room setting. Tests were performed in the same order on all participants.

### Participant Flow

All patients underwent primary rTSA with preoperative interscalene block as routine and were prescribed ondansetron, docusate, and hydrocodone/acetaminophen. Patients were routinely discharged on the day of surgery. A standardized postoperative physical therapy protocol was distributed to all participating physical therapists by the treating orthopedic surgeon. This protocol was intended to serve as a guideline rather than a directive, allowing physical therapists to individualize treatment based on each patient’s specific needs and clinical progress. Shoulder abduction with external and internal rotation was restricted for the first 12 weeks following surgery. Beginning two weeks postoperatively, patients were encouraged to initiate gradual, progressive active-assistive and active range of motion exercises within the prescribed limits. Sling immobilization was maintained continuously for the first four weeks, after which patients were instructed to discontinue sling use during the day but continue wearing it at night for an additional two weeks. Targeted deltoid strengthening exercises were introduced at approximately six weeks postoperatively.

## Predictive and Response Variables

### Domain 1: Patient Reported Outcome Measures

Patient-reported outcome measures (PROs) of pain and psychological function are biomarkers recommended for study in the acute-to-chronic pain signature initiative.^6^ PROs in the pain domain included primary biomarkers of general pain intensity, widespread body pain, and symptoms of neuropathic pain.^6^ General pain intensity was assessed using the 11-point Numerical Rating Scale (NRS-11), scored from 0 to 10, with 0 indicating no pain and 10 indicating the worst pain imaginable. The degree of widespread pain was assessed with the question “I feel pain all over my body.” This was scored on a five-point Likert scale with “Never” coded to 0, “Rarely” coded to 1, “Sometimes” coded to 2, “Often” coded to 3, and “Always” coded to 4. Self-reported pain duration (in months) was also recorded as a secondary biomarker.^6^ The degree of neuropathic pain was assessed using the painDETECT questionnaire.^12,13^ The painDETECT was developed by the German Research Network on Neuropathic Pain to establish a simple and validated screening tool to detect neuropathic pain components in patients with chronic lower back pain.^12^ The maximum possible score is 38, and the minimum possible score is −1, with a higher score indicating increased likelihood of neuropathic pain.

PROs in the psychological domain included the primary biomarkers of anxiety and depression. Anxiety was assessed with the Generalized Anxiety Disorder-7 (GAD-7).^14^ This is a 7-item self-report measure with a 4-point Likert scale for each item with a total score from 0 to 21. Higher scores indicate greater anxiety symptom severity. Depression severity was assessed with the Patient Health Questionnaire-9 (PHQ-9),^15^ which is widely used in outcome research of total joint arthroplasty.^16^ This is a 9-item self-report questionnaire with each item scored from 0 to 3, with higher scores indicating greater depression severity and likelihood of a depression diagnosis. All PRO measures were collected by a RedCap survey on a provided iPad.

### Domain 2: Patient Characteristics

Age, sex, race, ethnicity, education level, and relationship status were used as secondary biomarkers.^6^ These were collected verbally or by RedCap survey response on a provided iPad.

### Domain 3: Quantitative Sensory Testing

Quantitative sensory testing (QST) of the surgical site has been recommended as a primary biomarker, and of a control site as a secondary biomarker.^6^ Two recommended measures of QST are pressure pain threshold (PPT) and pinprick temporal summation of pain (TSP). Pain sensitivity can be measured with PPT algometry that assesses the least amount of sensory input required that is experienced as pain.^17^ Pressure algometry has shown to be reliable in evaluating tissue sensitivity.^18,19^ TSP can be generated by a variety of noxious stimuli including heat, pressure, or pin-prick.^20^ TSP represents the behavioral correlate of wind-up, a phenomenon of centralized facilitation where increased firing of spinal secondary neurons results from repeated C-fiber stimulation.^20,21^ Protocols vary for the assessment of TSP in people with musculoskeletal pain, however most published studies have used repeated nociceptive mechanical stimuli.^22^

Briefly, for both QST measures used in this study, participants were seated in a standard chair with feet on the floor and the shoulder supported by a table at 45º of abduction. Both PPT and TSP tests were of the mid-belly of the affected deltoid (surgical site), the unaffected deltoid (control site), and the unaffected side’s tibialis anterior (control site). At the shoulder, the site was marked at a site 3 cm distal to the acromion process to find the midpoint of the lateral deltoid muscle mass. At the tibialis anterior, the site was marked 4 cm distal and lateral to the tibial tuberosity. A Force One algometer (Wagner Instruments, Greenwich, CT) with the 0.875 cm^2^ tip was used for PPT, with the average of three values for each site recorded in kg/cm^2^. A lower value indicates greater sensitivity to pressure pain. A PinPrick stimulator (MRC Systems, Heidelberg, Germany) was used for one trial of TSP at each anatomical site. The difference between the peak pain experienced in 10 stimuli and the initial pain experienced was recorded as the temporal summation magnitude (TSMag).^23,24^ Before the first pin press, a 256 mN pin was presented. If this elicited pain (>0/10), this was used for the repeated tests. If this did not elicit pain, a 512 mN pin was used. A higher value TSMag indicates a greater pain facilitation response. An in-depth description of these testing procedures used for other shoulder conditions is available elsewhere.^24,25^

### Statistical Analysis

Statistical analyses were performed using SAS Version 9.4 (Cary, NC). Appropriate statistical assumptions were tested, and adjustments to models were made in cases of assumption violation. The 6-week change variables were assessed for distribution normality using the Shapiro-Wilk test. Given variable distributions varied across outcome measures, both a paired t-test of means and Wilcoxon signed-rank test of medians were used to identify statistically significant differences from preoperative to follow-up.

For the second objective, simple linear regression models were created for each independent continuous variable predictor with 6-week pain intensity improvement as the dependent variable. For sex, a Kruskal-Wallis test was used to compare the two groups. An α = 0.05 was used for the level of significance on all statistical tests. Missing data were not included in the simple linear regression models.

## Results

Twenty-one participants were enrolled. One participant did not attend the 6-week follow-up visit. One participant declined to undergo baseline temporal summation testing, and thus was not included in the analyses of temporal summation change. Two participants declined to complete all parts of the baseline PainDETECT, and thus were not included in the analyses of PainDETECT change. No others were missing data. The descriptive statistics of demographic and clinical characteristics are reported in Table I.

For the first objective, 6-week outcome measure changes are reported in Table II. Significant changes in group means and medians were found for shoulder pain intensity, widespread pain, and neuropathic pain symptoms. Specifically, patients reported a mean 3.6 (SD=4.7, p=0.0028) and median 4.5 (IQR=5.0, p=0.0023) improvement in pain intensity score from baseline to 6-weeks post-op. Reports of widespread pain also significantly improved, with a mean change score of 0.5 (SD=0.8, p=0.0248). Additionally, patients reported significant improvement with PainDETECT score, with a mean change score of 4.2 (SD=5.9, p=0.0077), and a median change of 5 (IQR=8, p=0.0071).

For the temporal summation magnitude of the surgical deltoid, there was a significant 6-week difference in mean 1.0 (SD=2, p=0.0462) but not median. One participant, at the 6-week visit, reported an initial pain report of 5 and a repeated pain report of 0. With the resulting TSMag score of −5, this was an outlier that affected the group mean. With repeating the test (n=18) without this participant included, neither test was statistically significant (t=-1.99, p=0.0611) and (S=-31, p=0.0844).

To investigate the associations between baseline factors and degree of pain improvement, the univariate regression models are reported in Table III. A significant association was found for baseline pain intensity (F=18.79, p=0.0004). This was a positive association with which participants with higher baseline pain can expect a greater improvement in pain. For each unit increase in preoperative pain level score, a 1.15 greater increase in pain improvement score can be expected (β=1.156, 95% CI 0.60 to 1.72). The R^2^ was 0.51 for this model. A significant association was found for baseline TSMag of the Tibialis Anterior (F=5.06, p=0.0380). An inverse association was observed: participants with higher baseline TSMag experienced smaller reductions in pain intensity. Specifically, for each one-unit increase in preoperative TSMag, the improvement in NRS-11 pain score was 1.17 points smaller (β = −1.17, 95% CI −2.26 to −0.07). The model explained 23% of the variance in pain reduction (R^2^ = 0.23).

## Discussion

We conducted a longitudinal cohort study to evaluate the 6-week changes in select biomarkers from before to after the rTSA procedure. We found significant changes in group means and medians for shoulder pain intensity, widespread pain, and neuropathic pain symptoms. A significant difference was found in the mean but not the median for surgical shoulder TSMag, which showed no difference after removing an outlier. Baseline shoulder pain level and TSMag of the tibialis anterior were the only significant baseline predictors for 6-week improvement in shoulder pain intensity. With two distinct anatomical sites providing significant associations, our results suggest future study of central sensitization, which will be explored for this cohort in a future manuscript.

### Main Findings and Clinical Application

Preoperatively, our cohort’s mean NRS-11 of 6.0 (SD=2.9) was nearly identical to a larger cohort who had a preoperative mean of 6.1 (SD 2.21). At a much later follow-up period (48.6±25.1 months), this cohort demonstrated a mean improvement of 5.0.^26^ In our sample’s short-term follow-up, shoulder pain intensity improved by a mean of 3.6 (t=3.43, p=0.0023) and a median of 4.5 points (S=72, p=0.0028), both greater than the reported minimum clinically important difference (MCID) of 1.4 and the substantial clinical benefit (SCB) of 2.6.^27^ Degree of improvement was most strongly predicted by baseline pain intensity, with higher baseline pain predicting greater improvement. This may reflect a ceiling effect, where those with lower pain at baseline have less room for improvement. This information can be used before surgery to help patients set realistic expectations regarding the magnitude of pain improvement 6 weeks following surgery.

### QST Changes from Baseline to 6 weeks

From baseline to follow-up, we found differing results between QST and self-reported measures of neuropathic pain. We found no statistically significant reduction in experimental evoked pain sensitivity (median TSMag) or PPT of any site. But with self-reported neuropathic pain symptoms, our participants reported a significant reduction in total PainDETECT score with a mean change of 4.2 and median change of 5. Although not yet established for rTSA, this is near the recommended MCID found with TKA of 5.0 for those with neuropathic pain and 3.0 for those with nociceptive pain.^28^ This identifies a discordance between outcome measures, as we had expected decreases in PainDETECT score to mirror decreases in experimental pain sensitivity.

We did not find baseline localized PPT to be associated with the magnitude of pain intensity improvement, which is similar to findings of THA or TKA outcomes.^9^ Higher baseline Tibialis Anterior TSMag predicted less shoulder pain improvement (β=-1.17, F=5.06, p=0.0380). With a higher TSMag of the TA, a marker of sensitivity at a distal anatomical site, a participant is more likely to demonstrate increased centralized facilitation of pain. This is in line with the results of research on control-site TS, with which TS at the contralateral middle phalanx was associated with less improvement three months following hip arthroscopy.^29^

### Limitations

It is difficult to evaluate postsurgical pain and chief complaint pain as separate constructs. We attempted to assess these as separate questions at the 6-week follow-up visit, but several of our participants expressed difficulty in discerning the difference. We could not follow the specific definition of chronic postsurgical pain at the 6-week visit due to the duration not yet reaching two months.^30^ The baseline data were not collected on the date of the surgery, and it is possible that participants underwent other interventions for pain between the time of the baseline data collection and the surgery. Because each participant’s surgery schedule differed, there was also a varying timeline between the time of baseline data collection and the time of surgery. Between surgery and the 6-week follow-up, analgesic medication regimens or additional supplementary treatments were not standardized as a requirement of study participation. Our sample size was not designed to provide high power to detect differences for each outcome measure, increasing the risk of Type I error.

### Future Research

In future studies with a larger sample size, multivariable modeling may be appropriate for an expansion of our second aim, with the goal to elucidate which baseline measures are the strongest individual predictors of surgical outcomes. Future studies of this procedure can include a more comprehensive set of biomarkers, a more in-depth, staged analysis of shoulder pain phenotypes,^31^ autonomic predictors, and the -omics and brain neuroimaging domains.^6^ Future studies can also include a longer follow-up period (6 months) that meets the published definition of chronic postsurgical pain.

## Conclusion

Several patient-reported outcome measures and experimental tests of pain sensitivity can serve as biomarkers of the short-term changes that can be expected following rTSA. Baseline pain intensity and temporal summation magnitude of the tibialis anterior were associated with the degree of pain improvement, suggesting their use in preoperative outcome prediction. Future research can include a larger sample size, a more comprehensive collection of biomarkers, and evaluate if these 6 week biomarker changes are associated with the development of chronic postoperative pain at 6 months.

## Supporting information

Table III

## Data Availability

All data produced in the present study are available upon reasonable request to the authors.

## Funding

This study and REDCap electronic data capture tools were supported in part by the UT Southwestern School of Health Professions Collaborative Research Grant Program. This was also supported in part by an Interdisciplinary Research Program grant from the University of Texas at Arlington. The use of REDCap in this publication was supported by the National Center for Advancing Translational Sciences of the National Institutes of Health under the CTSA Program award number 1UL1TR003163-04. The content is solely the responsibility of the authors and does not necessarily represent the official views of the NIH.

## Author Contributions

CJP contributed to study conceptualization, participant recruitment, data collection, data analysis, and manuscript writing and editing.

AJN contributed to participant recruitment, study conceptualization, data collection, and manuscript writing and editing.

CMA contributed to data analysis, analytic plan development, and manuscript writing.

BT contributed to participant recruitment, data collection, and manuscript writing and editing.

AC contributed to participant recruitment, data collection, and manuscript writing and editing.

TG contributed to manuscript writing, analytic plan development, and editing.

KB contributed to manuscript writing, analytic plan development, and editing.

RJ contributed to manuscript writing, analytic plan development, and editing.

JZ contributed to funding acquisition, study conceptualization, participant recruitment, data collection, data analysis, and manuscript writing and editing.

## Declaration of Generative AI and AI-assisted technologies in the writing process

During the preparation of this work the author(s) used Perplexity AI to identify grammatical and typographical errors. After using this tool/service, the author(s) reviewed and edited the content as needed and take full responsibility for the content of the publication.

## Disclaimer

None

This study was approved by the Institutional Review Board at the University of Texas (UT) Southwestern Medical Center (STU2021-0495).

## Note

Chris J. Pierson is now a postdoctoral research fellow at the University of Texas Medical Branch.

## Notes

### Competing Interest Statement

The authors have declared no competing interest.

