## Supplementary material for "Six-Week Changes in Pain Biomarkers Following Reverse Total Shoulder Arthroplasty: A Prospective Cohort Study": Table I

**Table I.** Participant demographic and clinical characteristics (n=20)

| **Sex, n, (%)** | |
| --- | --- |
| Male | 13 (65) |
| Female | 7 (35) |
| **Age, years** | |
| Mean (SD) | 67 (8) |
| Median (IQR) | 68 (6) |
| **Body Mass Index** | |
| Mean (SD) | 30.3 (4.8) |
| Median (IQR) | 30.7 (4.7) |
| **Shoulder pain duration, months** | |
| Mean (SD) | 98 (144) |
| Median (IQR) | 41 (98) |
| **American Shoulder and Elbow Surgeons Index Score (n=17)** | |
| Mean (SD) | 45.4 (22.6) |
| Median (IQR) | 43.4 (26) |
| **Race, n (%)** | |
| Black or African American | 2 (10) |
| White | 17 (85) |
| American Indian | 1 (5) |
| **Ethnicity, n (%)** | |
| Hispanic or Latino | 3 (15) |
| Non-Hispanic or Latino | 17 (85) |
| **Marital Status, n (%)** | |
| Married | 12 (60) |
| Widowed | 4 (20) |
| Divorced or Annulled | 3 (15) |
| Single/Never Married | 1 (5) |
| **Employment Status** | |
| Full Time | 4 (20) |
| Retired | 12 (60) |
| Disabled | 2 (10) |
| Not employed | 2 (10) |
| **Surgical Indication** | |
| Glenohumeral osteoarthritis | 4 (20) |
| Rotator cuff arthropathy | 16 (80) |
| **Operative Side, n (%)** | |
| Right | 10 (50) |

Note: N=21 were enrolled but n=20 were included in the final analysis.

Abbreviations: SD, standard deviation; IQR, interquartile range
