## Supplementary material for "Six-Week Changes in Pain Biomarkers Following Reverse Total Shoulder Arthroplasty: A Prospective Cohort Study": Table II

**Table 2.** Outcome measure changes (n=20)

|  | **Baseline** | | **Follow-Up** | **Change** | **Test Statistic** | **p-value** |
| --- | --- | --- | --- | --- | --- | --- |
| **Shoulder Pain Intensity** | | | | | | |
| Mean (SD) | 6.0 (2.9) | | 2.4 (3.3) | 3.6 (4.7) | t = 3.43 | **0.0028*** |
| Median (IQR) | 6 (3) | | 0 (4.5) | 4.5 (5.0) | S = 72 | **0.0023*** |
| **Widespread Pain (“I feel pain all over my body”)** | | | | | | |
| Mean (SD) | 1.1 (0.9) | | 0.6 (1.1) | 0.5 (0.8) | t = 2.43 | **0.0248*** |
| Median (IQR) | 1 (2) | | 0 (1) | 0 (1) | S = 23 | **0.0449*** |
| **GAD-7 Total Score** | | | | | | |
| Mean (SD) | 3.4 (4.3) | | 2.3 (3.8) | 1.1 (4.4) | t = 1.12 | 0.2762 |
| Median (IQR) | 2 (4.5) | | 0 (2) | 0 (2) | S = 18.5 | 0.2107 |
| **PHQ-9 Total Score** | | | | | | |
| Mean (SD) | 5.2 (4.4) | | 3.3 (2.8) | 1.9 (4) | t = 2.05 | 0.0541 |
| Median (IQR) | | 5 (7.5) | 2.5 (5) | 1 (5.5) | S = 31.5 | 0.1106 |
| **PainDETECT Total Score (n=18)** | | | | | | |
| Mean (SD) | | 8.1 (5.3) | 3.9 (4.5) | 4.2 (5.9) | t = 3.02 | **0.0077*** |
| Median (IQR) | | 6 (7) | 2.5 (4) | 5 (8) | S = 45.5 | **0.0071*** |
| **Surgical Deltoid TSMag (n=19)** | | | | | | |
| Mean (SD) | | 2.4 (2.3) | 1.5 (2.3) | 1.0 (2) | t = 2.14 | **0.0462*** |
| Median (IQR) | | 2 (4) | 2 (3) | -1 (2) | S = -34 | 0.0537 |
| **Opposite Deltoid TSMag (n=19)** | | | | | | |
| Mean (SD) | | 1.8 (2.4) | 2 (1.7) | -0.2 (2.5) | t = -0.28 | 0.7844 |
| Median (IQR) | | 1 (4) | 2 (2) | 1 (4) | S = -9.5 | 0.6612 |
| **Tibialis Anterior TSMag (n=19)** | | | | | | |
| Mean (SD) | | 1.6 (2.0) | 1.8 (1.6) | -0.2 (1.9) | t = -0.48 | 0.6354 |
| Median (IQR) | | 1 (3) | 1.5 (3) | 0 (1) | S = -5.5 | 0.6680 |
| **Surgical Deltoid PPT, kg/cm^2^** | | | | | | |
| Mean (SD) | | 3 (2) | 3.3 (1.7) | -0.3 (1.5) | t = -0.91 | 0.3750 |
| Median (IQR) | | 2.1 (4.1) | 3 (2.7) | -0.3 (1.1) | S = -34 | 0.2162 |
| **Opposite Deltoid PPT** | | | | | | |
| Mean (SD) | | 3.6 (2.3) | 3.3 (2) | 0.3 (1.6) | t = 0.91 | 0.3699 |
| Median (IQR) | | 3.3 (3.0) | 2.8 (2.6) | 0 (1.9) | S = 16 | 0.5706 |
| **Tibialis Anterior PPT** | | | | | | |
| Mean (SD) | | 4.4 (2.0) | 4.5 (2.2) | -0.1 (2.5) | t = -0.18 | 0.8555 |
| Median (IQR) | | 3.5 (3.5) | 3.8 (2.7) | -0.3 (1.6) | S = -9 | 0.7562 |

*p < 0.05

Abbreviations: SD, standard deviation; IQR, interquartile range; CSI, Central Sensitization Inventory; GAD-7, Generalized Anxiety Disorder-7; PHQ-9, Patient Health Questionnaire-9; TSMag, Temporal Summation Magnitude; PPT, Pressure Pain Threshold. One participant declined to undergo temporal summation testing. Two participants declined to complete all parts of the baseline PainDETCT. Change difference of baseline to follow up, paired t-test for mean comparison, Wilcoxon signed rank test S for median comparison
