## Supplementary material for "Six-Week Changes in Pain Biomarkers Following Reverse Total Shoulder Arthroplasty: A Prospective Cohort Study": Table III

**Table III.** Results of univariate predictive analyses

|  | **Shoulder Pain Intensity Improvement (n=20)** | | | | |
| --- | --- | --- | --- | --- | --- |
| **Baseline Variable** | | **Estimate (β)** | **Test Statistic** | **95% CI** | **p-value** |
| Sex | | - | X^2^ = 0.4080 | - | 0.5230 |
| Age | | -0.11 | F = 0.60 | -0.41-0.19 | 0.4478 |
| Pain Duration | | -0.00 | F = 0.41 | -0.02-0.01 | 0.5283 |
| Pain Intensity | | 1.16 | F = 18.79 | 0.60-1.72 | **0.0004*** |
| Widespread Pain (“I feel pain all over my body”) | | -2.31 | F = 4.26 | -4.67-0.04 | 0.0539 |
| GAD-7 Total Score | | 0.18 | F = 0.53 | -0.35-0.72 | 0.4772 |
| PHQ-9 Total Score | | 0.07 | F = 0.07 | -0.46-0.59 | 0.7899 |
| PainDETECT Total Score (n=18) | | 0.01 | F = 0.00 | -0.45-0.48 | 0.9503 |
| Surgical Deltoid TSMag (n=19) | | -0.29 | F = 0.34 | -1.33-0.75 | 0.5650 |
| Opposite Deltoid TSMag (n=19) | | -0.32 | F = 0.44 | -1.33-0.69 | 0.5155 |
| Tibialis Anterior TSMag (n=19) | | -1.17 | F = 5.06 | -2.26 to -0.07 | **0.0380*** |
| Surgical Deltoid PPT, kg/cm^2^ | | -0.14 | F = 0.07 | -1.28-1.0 | 0.7977 |
| Opposite Deltoid PPT, kg/cm^2^ | | -0.55 | F = 1.39 | -1.54-0.43 | 0.2537 |
| Tibialis Anterior PPT, kg/cm^2^ | | 0.10 | F = 0.03 | -1.08-1.29 | 0.8567 |

*p-value < .05

Abbreviations: CI, confidence interval; CSI, Central Sensitization Inventory; GAD-7, Generalized Anxiety Disorder-7; PHQ-9, Patient Health Questionnaire-9; TSMag, Temporal Summation Magnitude; PPT, Pressure Pain Threshold
